# Self-testing for proteinuria in pregnancy: a systematic review and meta-analysis

**DOI:** 10.1101/2022.08.13.22278735

**Authors:** Ping Teresa Yeh, Dong Keun Rhee, Caitlin Elizabeth Kennedy, Chloe A. Zera, Özge Tunçalp, Briana Lucido, Manjulaa Narasimhan

## Abstract

**Background:** Self-testing for proteinuria may help identify preeclampsia risk during pregnancy, increase end-user empowerment, and reduce burden on health systems. We conducted this systematic review of the impact of proteinuria self-testing during pregnancy to expand the evidence base for the World Health Organization consolidated guideline on self-care interventions.

**Methods:** We comprehensively searched for articles comparing the effect of proteinuria self-testing with clinic-based testing among pregnant individuals receiving antenatal care on the following outcomes: maternal mortality or near-miss; adverse pregnancy outcomes; eclampsia or pre-eclampsia; long-term diseases; follow-up care and appropriate management; self-efficacy, self-determination, autonomy, and empowerment; mental health and well-being; adverse events and social harms; device-related issues; intra-uterine growth restriction; preterm birth; and stillbirth or perinatal death. After abstract screening and full-text review, we systematically extracted data using standardized forms and summarized the relative risks of outcomes between self-testing and clinic-based testing for proteinuria. We also assessed values and preferences and costs of self-testing.

**Results:** Three publications were identified for the effectiveness review; two for values/preferences, and none for the cost review, mostly from high-income countries. Overall, there was no statistically significant difference between self-testing and inpatient care for proteinuria among women with hypertension for any of the outcomes with data available. In general, both women and providers were supportive of proteinuria self-testing because it allows a greater role in self-care and fewer clinic visits, though some emphasized the need to train end-users for proper testing and appropriate follow-up actions.

**Conclusions:** Very limited evidence suggests that self-testing for proteinuria results in comparable maternal and fetal outcomes as provider-testing for hypertensive pregnant individuals, and is generally acceptable to end-users and providers despite some concerns. No evidence of effectiveness was available for the general pregnant population. This evidence base supports its feasibility as an additional option for identifying individuals at risk of preeclampsia.

**Systematic review registration number:** PROSPERO CRD42021233845

## Background

Preeclampsia is a significant cause of maternal and perinatal morbidity and mortality, affecting 2-8% of pregnancies worldwide.[1] This complication is generally diagnosed in pregnant individuals who experience an onset of hypertension and subsequent proteinuria (greater than 300 mg in one day of protein in urine) during pregnancy.[2] About one-third of individuals who experience onset of proteinuria past 20 weeks of pregnancy may ultimately contract preeclampsia and have increased risk of adverse pregnancy and birth outcomes.[3, 4] Therefore, monitoring urine protein levels during pregnancy serves as an important intervention in achieving early diagnosis and care for preeclampsia in pregnant individuals.

Measuring proteinuria early in pregnancy can help identify individuals who are at a high risk of preeclampsia and related complications, including preterm delivery and fetal malformations.[5] Screening for proteinuria is typically done through dipstick urinalysis, which requires a small sample of clean urine and provides a rapid result.[6] Since a recognized limitation of dipstick urinalysis is poor specificity for preeclampsia versus kidney function,[7] subsequent 24-hour urine collection or spot urine protein:creatinine ratio is used to verify positive proteinuria findings from dipstick testing [8] as clinically appropriate. Dipstick urinalysis is typically done at the point-of-care during routine antenatal care (ANC) contacts. Yet, frequency of ANC contacts and proteinuria screening varies by setting, and disparities exist between settings.[9] Routine screening in clinic is time-consuming and can become expensive due to frequent false positives, which require further testing.[6]

Emerging research suggests that screening can also be done through self-testing. Self-testing of proteinuria may be useful for early detection and care of preeclampsia, and for reducing the burden of care visits during pregnancy. Self-testing may also help pregnant individuals feel involved with their care. Self-testing may also be a feasible way to promote health awareness and management during health emergencies like the COVID-19 pandemic, when access to healthcare services may be limited; a pulse survey conducted by the World Health Organization (WHO) on the continuity of essential health services during the COVID-19 pandemic showed that ANC services were among the most severely disrupted.[10] We conducted this systematic review in the context of expanding the evidence base of the WHO’s guideline on self-care interventions [11] to include additional considerations related to maternal and perinatal health.

## Methods

In this review, we evaluated the current literature to inform decisions on whether self-testing for proteinuria during pregnancy should be available in addition to clinic-based testing. We assessed three areas relevant to this topic: (1) effectiveness of the intervention, (2) values and preferences of end users and providers, and (3) cost information. We followed the Preferred Reporting Items for Systematic review and Meta-Analysis (PRISMA) guidelines,[12] and the protocol was registered on the International Prospective Register of Systematic Reviews (PROSPERO #CRD42021233845). Ethical approval was not required for this systematic review, since all data came from published articles.

### Effectiveness review inclusion criteria

The effectiveness review was designed according to the PICO (Population, Intervention, Comparison, Outcomes) format as follows:

– **Population:** Pregnant individuals
– **Intervention:** Self-testing for proteinuria (either by the pregnant individual or by another layperson, such as a family member)
– **Comparison:** Clinic proteinuria testing by health care providers during ANC contacts only
– **Outcomes:**
  - Maternal outcomes:
    1. Maternal mortality or near-miss
    2. Adverse pregnancy outcomes (e.g. spontaneous abortion, premature rupture of membrane, placental abruption)
    3. Eclampsia or pre-eclampsia
    4. Long-term (after pregnancy) cardiovascular risk, chronic hypertension, diabetes, stroke
    5. Follow-up care and appropriate management
    6. Self-efficacy, self-determination, autonomy, and empowerment
    7. Mental health and well-being (anxiety, stress, self-harm)
    8. Adverse events and social harms (including discrimination, intimate partner violence, stigma), and whether these harms were corrected/had redress available
    9. Device-related issues (e.g. test failure, problems with manufacturing, packaging, labeling, or instructions for use)
  - Fetal/newborn outcomes:
    1. Intra-uterine growth restriction
    2. Preterm birth
    3. Stillbirth or perinatal death

For our review, we included studies that compared self-testing for proteinuria to the comparator group. The studies could be randomized controlled trials (RCTs), non-RCTs, or comparative observational studies, which would include prospective controlled cohort studies, cross-sectional studies, controlled before-after studies and interrupted time series, as long as they compared individuals who received the intervention to those who did not. In addition, the included studies were limited to peer-reviewed publications that measured one or more outcomes of interest to our review.

No restrictions were placed based on the location of intervention. No language restrictions were used on the search. Articles in English, French, Spanish, and Chinese were coded directly; articles in other languages were translated.

### Search strategy

We searched PubMed, Cumulative Index to Nursing and Allied Health Literature (CINAHL), Latin American & Caribbean Health Sciences Literature (LILACS) and Embase through the search date of November 16, 2020 using the search string as follows (designed for PubMed and adapted for other databases):

(proteinuria [Mesh] OR proteinuria [tiab] OR “urinary protein excretion” [tiab] OR “urinalysis” [Mesh] OR “reagent strips” [Mesh] OR creatinine [Mesh] OR “dipstick”[tiab])

AND

(pregnancy [Mesh] OR pregnancy [tiab] OR pregnant [tiab] OR peri-natal [tiab] OR perinatal [tiab] OR antenatal [tiab] OR maternal [tiab])

AND

(“self care”[Mesh] OR “self-care”[tiab] OR “self-monitoring”[tiab] OR “self-management”[tiab] OR “self-monitor”[tiab] OR “self-manage”[tiab] OR “self-monitored”[tiab] OR “self-managed”[tiab] OR “self-evaluate”[tiab] OR “self-evaluating”[tiab] OR “self-evaluation”[tiab] OR “self-test”[tiab] OR “self-testing”[tiab] OR “home”[tiab] OR “pharmacy”[tiab])

We also searched for ongoing RCTs through clinicaltrials.gov, the WHO International Clinical Trials Registry Platform, the Pan African Clinical Trials Registry, and the Australian New Zealand Clinical Trials Registry. Additionally, we searched the Cochrane Library for primary research articles cited in relevant reviews. Secondary reference searching was also conducted on all studies included in this review, and we asked experts in the field to help us identify additional articles.

A member of the study staff screened the titles, abstracts, citation information, and descriptor terms of citations that were identified through the search strategy. Full text articles were obtained of all abstracts, and two independent reviewers assessed them for final study selection. Differences were resolved through consensus and, if needed, with the intervention of senior staff members.

### Data extraction, management, and analysis

Data were extracted independently by two reviewers using standardized data extraction forms. Differences in data extraction were resolved through consensus and referral to a senior study team member from WHO when necessary. The components of information that were gathered included: (1) study identification (authors, type of citation; year of publication), (2) study description (study objectives, location, population characteristics, type of proteinuria monitoring such as the brand of urine dipstick), (3) description of self-testing access, (4) description of any additional intervention components (e.g. any education, training, support provided), (5) study design, (6) sample size, (7) follow-up periods, (8) loss to follow-up, and (9) outcomes (analytic approach, outcome measures, comparison groups, effect sizes, confidence intervals, significance levels, conclusions, study limitations).

Data were analyzed according to coding categories and outcomes. For each outcome assessed in the review, we summarized data in Grading of Recommendations Assessment, Development and Evaluation (GRADE) Evidence Profile tables using GRADEPro [15] and in a summary of effects table. When multiple studies reported the same outcome, meta-analysis was conducted with program Comprehensive Meta-Analysis (CMA) using random-effects models to combine risk ratios. Heterogeneity was assessed using I-squared and Q statistics. We used RCT data where they were available; if RCT data were not available for an outcome, we would have pulled data from observational studies. For RCTs, risk of bias was assessed using the Cochrane Collaboration’s tool for assessing risk of bias.[13] For non-RCTs but comparative studies, study rigor would have been assessed using the Evidence Project 8-item checklist for intervention evaluations.[14]

Where possible, all analyses were stratified by the following categories or subgroups: (1) location or context of self-testing (ambulatory, hospitalized, or additional to standard antenatal clinic contacts), (2) prior risk of preeclampsia, (3) vulnerabilities (e.g. obesity, age, poverty, disability, rural/urban, literacy/educational level), and (4) high-income versus low- or middle-income countries.

### Complementary reviews

The same search terms were used to search and screen for studies to be included in the values and preferences and costs reviews. These studies could be qualitative or quantitative in nature, but had to present primary data collection – think pieces and review articles were not included. Literature was summarized qualitatively and organized by study design and methodology, location, and population.

#### Values and preferences review

Studies were included in this review if they presented primary data examining preferences of individuals regarding self-testing of proteinuria during pregnancy. We focused on studies examining the values and preferences of individuals who were self-testing for proteinuria during pregnancy or potentially eligible for this intervention. We also included studies examining the values and preferences of healthcare providers. We considered issues related to eligibility, accessibility, informed decision-making, coercion, and seeking redress in this section; this included the effects of stock-outs or availability of urine dipsticks.

#### Cost review

Studies were included in this review if they presented primary data comparing costing, cost-effectiveness, cost-utility, or cost-benefit of the intervention and comparison listed in the PICO question above, or if they presented cost-effectiveness of the intervention as it related to the PICO outcomes listed above. This included both cost to the health system and cost to the end-user. Cost literature was classified into four categories: health sector costs, other sector costs, patient/family costs, and productivity impacts.

### Patient and public involvement

Feedback on the review protocol and analysis was received from the WHO patient safety working group. Patients were involved in a global survey of values and preferences conducted to inform the WHO guideline on self-care interventions and play a role in the overall recommendation informed by this review.

## Results

Our database search yielded 398 records, and we identified another 8 through hand- and secondary searching (Figure 1). Of the 334 unique records, we retained 20 for full-text review. Ultimately, we included three studies in the effectiveness review, two in the values and preferences review, and none in the cost review.

**Figure 1.**
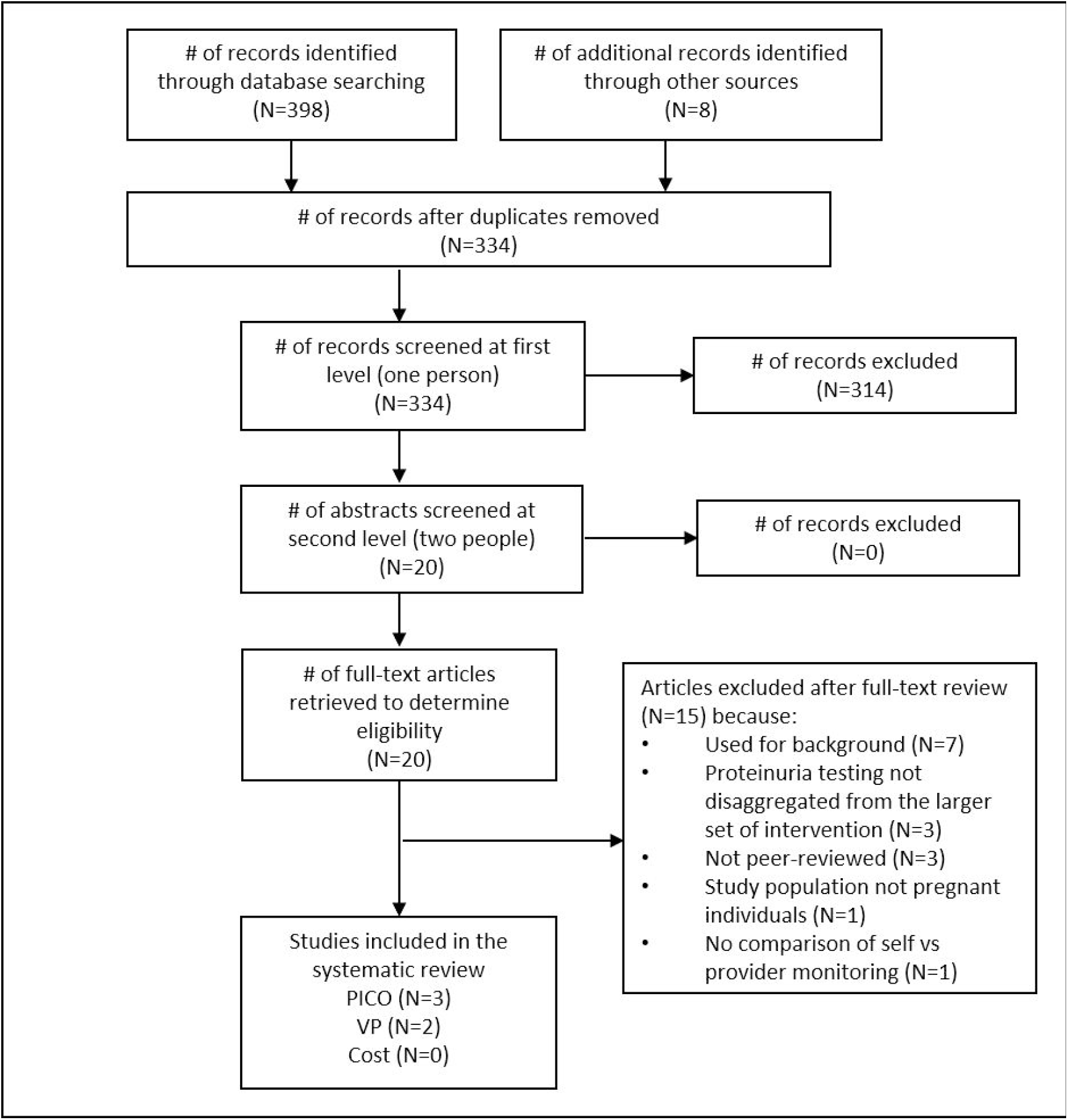
PRISMA flow chart showing disposition of citations through the search and screening process.

### Effectiveness review

Three studies met the inclusion criteria for the effectiveness review. These were small RCTs (sample sizes ranged from n=63 to n=218) from the United Kingdom (UK), Zimbabwe, and Hong Kong taking place over 20 years ago, comparing the effectiveness of antenatal ward admission (including proteinuria testing at the hospital) versus home-based standard of care (including proteinuria self-testing using dipsticks) among pregnant participants with varying degrees of non-proteinuric hypertension.[15-17] In the control (self-testing) arm of the included studies, which is the intervention for this review, pregnant women were given instructions on how to self-test for proteinuria, were provided an instruction sheet and container of urine dip sticks to take home, and were encouraged to test every day. They were advised to continue normal activity at home with no particular restrictions and received regular ANC at the local day care center or outpatient clinic either weekly [15, 16] or once every two weeks [17]. These participants were given instructions to contact the health facility for a consult or to be admitted if the dipsticks indicated proteinuria, among other health conditions (e.g. other signs/symptoms of pre-eclampsia like increasing headache or abdominal pain, indicators of labor, no fetal movement). In the intervention arm of the included trials, pregnant participants were admitted to the antenatal ward after study recruitment and received daily examination by health providers (e.g. obstetricians, midwives), blood pressure monitoring, and urine proteinuria testing. Activity (while an inpatient or on bed rest) was limited, though voluntary ambulation (e.g. to meals and toilet) was allowed.

For the purposes of our review, the relative risks of outcomes were recalculated to treat self-testing at home as the intervention and clinic/hospital-based testing as the control. Study descriptions are presented in Table 1 and the summary of effects in Table 2. It was not possible to further stratify the outcome data given the small number of studies. All RCTs reported on the development of maternal severe hypertension, birthweight in grams, small-for-gestational age, and preterm birth. Low birthweight was reported in the Zimbabwe study and Hong Kong study.[15, 16] The UK study reported on eclampsia, albuminuria, stillbirth, and neonatal mortality;[17] the Hong Kong study reported on maternal development of proteinuria and development of severe proteinuria.[16] In general, the certainty of evidence for the reported outcomes was low to very low because of indirectness (the comparator – hospital admission – went beyond our desired clinic-based proteinuria testing during ANC, and the intervention – home-based normal activity with daily proteinuria testing – included more components than solely proteinuria self-testing) and imprecision (very small sample sizes and very low event rates).

**Table 1.** Description of included studies.

| **Study** | **Location** | **Population** | **Sampling** | **Intervention** | **Comparator** |
| --- | --- | --- | --- | --- | --- |
| Mathews 1977<br>RCT | United Kingdom: Kent<br>Rural | Pregnant women (>28 weeks gestation, singleton) with diastolic blood pressure 90-109 mm Hg after 5 minutes' rest, without sedation<br>N=28 (self); 35 (provider) | Non-probability facility-based | Daily self-testing for urine proteinuria testing at home via Albustix, as part of "routine outpatient care" (reviewed biweekly until 36 weeks, every week thereafter) and "normal activity at home" | Proteinuria testing by the nurse/physician at the clinic, as part of "inpatient bed rest in hospital without sedation" |
| Crowther 1992<br>RCT | Zimbabwe: Harare<br>Urban | Pregnant women (28-38 weeks gestation, singleton) with diastolic blood pressure 90-110 mm Hg but no/trace proteinuria<br>N=108 (self); 110 (provider) | Non-probability facility-based | Daily self-testing for urine proteinuria at home via Albustix, as part of "routine outpatient care" (reviewed weekly) and "normal activity at home" | Proteinuria testing on a daily basis by the nurse/physician at the clinic, as part of "admission to hospital for bed rest" |
| Leung 1998<br>RCT | Hong Kong<br>Urban | Pregnant women (28-38 weeks gestation, singleton) with diastolic blood pressure 90-100 mm Hg after 5 minutes' rest<br>N=44 (self); 44 (provider) | Non-probability facility-based | Daily self-testing urine proteinuria at home via Albustix, as part of routine care at a day care clinic or outpatient care and "normal activity at home" | Proteinuria testing on a daily basis by the nurse/physician at the clinic, as part of "inpatient admission to antenatal ward for bed rest" |

**Table 2.** Summary of effects from meta-analyses (number of studies n>1) or from single studies.

| Outcome | Study type | Number of studies | RR | 95% CI | p-value for RR | Q-value | p-value for Q statistic | I-squared |
| --- | --- | --- | --- | --- | --- | --- | --- | --- |
| Maternal Outcomes |  |  |  |  |  |  |  |  |
| *Eclampsia or pre-eclampsia* |  |  |  |  |  |  |  |  |
| Eclampsia | RCT | 1 | No events reported in either arm |  |  |  |  |  |
| *Long-term cardiovascular risk, chronic hypertension, diabetes, stroke* |  |  |  |  |  |  |  |  |
| Albuminuria | RCT | 1 | 0.63 | 0.12 to 3.17 | 0.57 | NA | NA | NA |
| Development of severe hypertension (assessed with: diastolic blood pressure >109 mm Hg) | RCT | 3 | 0.91 | 0.30 to 2.70 | 0.86 | 5.28 | 0.07 | 62.10 |
| Development of proteinuric hypertension and DBP < 110 mm Hg | RCT | 1 | 1.29 | 0.23 to 7.24 | 0.77 | NA | NA | NA |
| Development of proteinuria (assessed with: >1+ on albustix testing) | RCT | 1 | 1.02 | 0.83 to 1.25 | 0.86 | NA | NA | NA |
| Development of severe proteinuria (assessed with: >3+ on albustix testing) | RCT | 1 | 1.44 | 0.92 to 2.26 | 0.11 | NA | NA | NA |
| Fetal/Newborn Outcomes |  |  |  |  |  |  |  |  |
| *Intrauterine growth restriction* |  |  |  |  |  |  |  |  |
| Birthweight in grams (difference in means) | RCT | 3 | -20.96 | -134.51 to 92.58 | 0.72 | 1.29 | 0.53 | 0 |
| Small for gestational age (assessed with: birthweight <10th percentile) | RCT | 3 | 0.91 | 0.52 to 1.58 | 0.74 | 1.97 | 0.37 | 0 |
| Low birthweight (assessed with: birthweight <2500 grams) | RCT | 2 | 1.52 | 0.79 to 2.93 | 0.21 | 0.03 | 0.87 | 0 |
| *Preterm birth* |  |  |  |  |  |  |  |  |
| Preterm birth (assessed with: delivery at gestational age <37 weeks) | RCT | 3 | 1.66 | 0.95 to 2.91 | 0.08 | 1.15 | 0.56 | 0 |
| *Stillbirth or perinatal death* |  |  |  |  |  |  |  |  |
| Stillbirth (assessed with: fetal death before onset of labor) | RCT | 1 | No events reported in either arm |  |  |  |  |  |
| Neonatal mortality (assessed with: number of deaths at 0-28 days of birth) | RCT | 1 | No events reported in either arm |  |  |  |  |  |

#### Eclampsia or pre-eclampsia

Though one RCT measured eclampsia as an outcome, no events of eclampsia were reported in either study arm.[17] This was graded as very low certainty evidence that self-testing for proteinuria had no difference compared to clinic-based testing on eclampsia or pre-eclampsia.

#### Long-term cardiovascular risk, chronic hypertension, diabetes, stroke

Three RCTs included indirect measures of long-term cardiovascular risk, chronic hypertension, diabetes, and stroke.[15-17] Meta-analysis of three RCTs found no difference in the risk of developing severe hypertension between pregnant women who self-tested for proteinuria and those who tested in clinics (RR: 0.91, 95% CI: 0.30-2.70, I-squared: 62.10). Between pregnant women who self-tested for albuminuria and those who were tested at a clinic, one RCT found no statistically significant difference in the risk of developing albuminuria (RR: 0.63, 95% CI: 0.12-3.17).[17] One RCT found no difference in the risk of developing proteinuric hypertension when comparing pregnant women who self-tested for proteinuria to pregnant women at the clinic (RR: 1.29, 95% CI: 0.23-7.24).[16] The same RCT found no difference between the self-testing and clinic-based testing arms in the risk of the pregnant women developing proteinuria (RR: 1.02, 95% CI: 0.83-1.25) or severe proteinuria (RR: 1.44, 95% CI: 0.92-2.26).[16] These outcomes were graded as low to very low certainty evidence showing that self-testing for proteinuria is comparable to clinic-based testing on long-term maternal health outcomes.

#### Intra-uterine growth restriction

The outcome of intra-uterine growth restriction was measured indirectly through three reported neonatal outcomes: birthweight in grams, low birthweight, and small for gestational age. Meta-analysis of three RCTs found no difference in birthweight between self-testing and clinic-based testing for proteinuria (MD: -20.96g, 95% CI: -134.51-92.58, I-squared: 0).[15-17] Meta-analysis of two RCTs showed no impact of proteinuria self-testing on the risk of low birthweight (1.52, 95% CI: 0.79-2.93, I-squared: 0).[15, 16] Meta-analysis of three RCTs found no difference in the risk of infants being born small-for-gestational-age (RR: 0.91, 95% CI: 0.52-1.58, I-squared: 0).[15-17] This was graded as low to very low certainty evidence that self-testing for proteinuria had no difference from clinic-based testing on intra-uterine growth restriction.

#### Preterm birth

Meta-analysis of three RCTs found an elevated but not statistically significant rate of preterm birth among women who self-tested for proteinuria compared to those who tested at the clinic (RR: 1.66, 95% CI: 0.95-2.91, I-squared: 0).[15-17] This was graded as very low certainty evidence that self-testing for proteinuria had no difference compared to clinic-based testing on preterm birth.

#### Stillbirth or perinatal death

One RCT measured stillbirth and neonatal mortality as outcomes; however, no events for either outcome were reported in either study arm.[17] This was graded as low to very low certainty evidence that self-testing for proteinuria had no difference compared to clinic-based testing on stillbirth or perinatal death.

#### Other outcomes of interest

Within the included studies, no comparative data were found for the following outcomes: maternal mortality or near-miss; adverse pregnancy outcomes; follow-up care and appropriate management; self-efficacy, self-determination, autonomy, and empowerment; mental health and well-being; adverse events and social harms, and whether these harms were corrected/had redress available; or device-related issues.

### Values and Preferences Review

Two quantitative feasibility studies for self-testing urine for proteinuria during pregnancy, one from the UK [18] and the other from the United States [19], found that most pregnant individuals were highly satisfied with or preferred self-testing for proteinuria over in-clinic testing. The common reason for liking self-testing for proteinuria across the two studies was ease of use.

One study explored values and preferences of end-users and providers in greater depth.[18] In this study, at least 95% of the surveyed women expressed willingness to self-test or discuss the results with their providers. All agreed that self-testing would provide them with a sense of greater involvement in their pregnancy care, and many appreciated the reassurance they felt when they received negative test results. Although some concerns were raised about increased stress or anxiety and accurate use of the dipsticks, almost all of the women who had not self-tested previously were open to the idea if provided the necessary training, reassurance, or second opinions from providers. These concerns were generally lower amongst the survey participants who had previously self-tested for proteinuria. In addition, these individuals felt that self-testing was empowering and they liked not making unnecessary trips to the hospital.

The majority of surveyed providers saw self-testing for proteinuria as a way for women to detect pre-eclampsia early, empower themselves, and save time and money.[18] Close to 80% believed that self-testing would enhance usual care, though about 70% also reported that they would repeat urinalysis despite having women self-test. However, providers also raised concerns about pregnant individuals’ aptitude and suitability for self-testing, their ability to act appropriately on any positive results, and whether self-testing might increase demand for urgent clinic-based services.

### Cost Review

No studies were identified for the cost review.

## Discussion

Among pregnant individuals with varying levels of non-proteinuric hypertension, three RCTs found no difference between self-testing and clinic-based testing for proteinuria on the risk of any of the maternal or neonatal outcomes for which data were available. These trials focused on individuals with an existing diagnosis of hypertension and compared home versus hospital management; we were unable to find any trials that compared self-testing to clinic-based testing for proteinuria during ANC contacts. We also found no comparative data for our other maternal outcomes of interest. In terms of values and preferences, most individuals found self-testing for proteinuria acceptable. Though some had concerns over their ability to accurately perform and interpret the tests, these fears were generally attenuated if training was provided. The sense of self-empowerment, ownership of care, and decreased frequency of clinic visits were also appreciated by end-users. Self-testing was positively regarded by providers for similar reasons. Providers generally approved adding proteinuria self-testing as a supplemental option to usual ANC, despite some concerns about end-users’ ability to perform the tests correctly or interpret/follow-up appropriately on the test results. There was no data available regarding cost savings related to self-testing or clinic-based testing, either for individuals or health system. However, even where urine testing strips are available over-the-counter through pharmacies or recommended by health care providers, their cost may not be covered by health insurance and may need to be out-of-pocket for many pregnant individuals. Cost is therefore an important consideration for an intervention which may need to be conducted over several months during pregnancy.

Several studies have examined the validity of self-testing for proteinuria, compared with clinic-based testing. A prospective observational study from Australia found that pregnant individuals who self-tested for proteinuria with dipstick tended to overestimate protein levels in their urine.[20] Another prospective cohort study found that the dipstick test performance was similar to the other proteinuria assessment methods that are typically used in the clinic setting, including protein:creatine ratio and 24-hour collection checks.[21]

Even so, controversy surrounding the clinical utility of urine dipstick testing (including self-testing) persists, in part due to the lack of a gold standard for diagnosis of proteinuria in pregnancy. Urine dipstick as an indicator for proteinuria is subject to several limitations, including variability in urine concentration depending on fluid status, the time of the day during which the test takes place, and whether they had urinated prior to testing. A diagnosis of proteinuria may require, at minimum, the indicator value of +2 on a dipstick, which should still be confirmed by a quantitative test.[22] Considering this lack of specificity, many providers prefer using spot urine protein-to-creatinine ratio for proteinuria testing because of its relatively high accuracy, reliability, and reproducibility.[23] Spot urine testing may be inferior to a 24-hour urine collection test due to the same limitations of dipstick urinalysis: high within-participant variability in urine protein excretion even among providers.[24] However, 24-hour urine collection is limited by the risk of contamination, over-collection, and inconvenience.

Despite recognized limitations, initial proteinuria testing (whether in clinic or at home) based on dipstick urinalysis with follow-up tests as indicated may help appropriately triage patients in resource-limited settings,[25] though the value-added of routine proteinuria testing via urine dipsticks may be limited in well-resourced settings. This testing method remains a standard tool for proteinuria testing in low-resource settings where testing affordability is a key issue. With the wide variety of commercially available dipsticks, there are some affordable and effective options.[26, 27] In low-resource settings, clinic-administered urinalysis which requires access to laboratory resources is more limited than other methods to screen for preeclampsia such as blood pressure monitoring,[27] suggesting that self-testing for proteinuria may still have value to help guide appropriate level of care. Further studies will be needed to adequately assess this question.

Clinical guidelines [28] highlight the need for information beyond proteinuria to diagnose and manage hypertensive complications of pregnancy, as non-proteinuric hypertensive disease is a recognized entity that has outcomes similar to preeclampsia. Though measuring proteinuria early in pregnancy can help predict individuals at high risk for important complications like preeclampsia and preterm delivery,[5] additional tests are needed for accurate assessment of maternal health status. Other indicators of preeclampsia including blood pressure, glomerular filtration rate, and neurological signs may be more important than proteinuria in predicting adverse pregnancy outcomes.[2, 29, 30]

This review has several strengths. We conducted a comprehensive screen across multiple databases as well as a hand search and secondary search, leaving little room for missing any relevant articles. We also examined the methodological quality of studies and assessed not only the effectiveness of self-testing for proteinuria but also the values and preferences of the pregnant end-users and providers. We found that proteinuria self-testing among pregnant individuals with diagnosed hypertension may be just as effective as proteinuria testing during inpatient care in predicting a wide range of maternal and neonatal outcomes, and that there is high acceptance of self-testing among both the pregnant individuals and providers who were surveyed.

It is important to interpret our findings within the context of limited literature. The studies identified for the effectiveness review were conducted among pregnant individuals diagnosed with some form of hypertension, which is one of the risk factors for preeclampsia and eclampsia, not the broader pregnant population. These effectiveness studies compared self-testing to inpatient admission, which is more intensive and less comparable than clinic-based testing. While we meta-analyzed the outcome of severe hypertension, we integrated only three small RCTs, so the meta-analysis may provide a false sense of certainty that the pooled estimate represents the true effect. Because these three studies were conducted decades ago, it may be challenging to apply their findings in current clinical practice. The scope of this review was limited to self-testing for proteinuria rather than any other of the many methods or biomarkers for identifying pregnancy complications, resulting in a small pool of studies meeting our inclusion criteria. We also found no peer-reviewed evidence on the costs related to proteinuria self-testing, although dipstick testing is likely substantially cheaper than hospitalization. Nonetheless, the currently available albeit limited data suggests that self-testing for proteinuria is not harmful. Further studies will be needed to assess whether self-testing for proteinuria as part of routine antenatal care can improve pregnancy outcomes among the general population, considering factors such as previous history of preeclampsia or eclampsia, age, obesity, race/ethnicity, and multiple pregnancies.

## Conclusions

Very limited evidence suggests that self-testing for proteinuria yields comparable maternal and fetal outcomes as provider testing, and is generally acceptable to end-users and providers. This evidence supports its feasibility as an additional option for identifying individuals at risk of preeclampsia.

## Data Availability

All data generated or analysed during this study are included in this published article and come from other published articles cited as included studies in this review.

## List of abbreviations

ANC: antenatal care
CINAHL: Cumulative Index to Nursing and Allied Health Literature
CMA: Comprehensive Meta-Analysis
GRADE: Grading of Recommendations Assessment, Development and Evaluation
LILACS: Latin American & Caribbean Health Sciences Literature
PICO: Population, Intervention, Comparison, Outcomes
PRISMA: Preferred Reporting Items for Systematic review and Meta-Analysis
PROSPERO: Prospective Register of Systematic Reviews
RCT: randomized controlled trial
UK: United Kingdom
WHO: World Health Organization

## Declarations

### Ethics approval and consent to participate

Not applicable.

### Consent for publication

Not applicable.

## Authors’ contributions

MN conceptualized the study following input from OT. CEK and PTY designed the protocol with feedback from OT, BL, and MN. PTY ran the database search and oversaw search, screening, full text review, and data abstraction processes with support from DR. CEK and PTY performed data analysis. PTY and DR drafted the manuscript. PTY, DR, CEK, CAZ, OT, BL, and MN reviewed the draft, provided critical review, and read and approved the final manuscript. The corresponding author, as guarantor, accepts full responsibility for the finished article has access to any data and controlled the decision to publish. The corresponding author attests that all listed authors meet the authorship criteria and that no others meeting the criteria have been omitted. The named authors alone are responsible for the views expressed in this publication and do not necessarily represent the decisions or the policies of the World Health Organization (WHO) nor the UNDP-UNFPA-UNICEF-WHO-World Bank Special Programme of Research, Development and Research Training in Human Reproduction (HRP).

## Acknowledgements

We thank Maurice Bucagu, Laura Ferguson, Rudolfo Gomez, Oleg Kuzmenko, and Karima Gholbzouri for their feedback on the review protocol. We also thank our Johns Hopkins graduate research assistants (Huneid Kautsar, Jaime Marquis, and Cynthia Li) for their crucial help in searching, screening, and extracting data.

